# “A Population-Specific Breast Cancer Risk Prediction Model for Indian Women (A Pilot Study): Advancing Beyond Traditional Assessment Tools”

**DOI:** 10.1101/2025.07.20.25331883

**Authors:** Prasoon Prakash, Kanika Arora, Arushi Gupta, Eshita Zjigyasu, Vishal Vivek Saley, Vipul Kumar Rathore, Aarjav Satia, Chetan Arora, Mausam, Krithika Rangarajan, Amit Gupta, Sarthak Singh, Hari Sagiraju, Kalpa Jyoti Das, Jitendra Kumar Meena, Ishaan Gupta

## Abstract

**Background:** Breast cancer is the most prevalent cancer among women in India, characterized by late-stage diagnoses and high mortality rates. Existing breast cancer risk prediction models, such as the Gail and Tyrer-Cuzick models, were primarily developed using Western datasets, limiting their applicability to the Indian context due to socio-demographic, genetic, and cultural differences.

**Objective:** This pilot study aims to develop and validate a machine learning (ML)–based breast cancer risk prediction model tailored specifically to the Indian population, addressing the limitations of traditional tools, with the potential for future methodological expansion to build more robust and generalizable models.

**Methods:** A retrospective case-control pilot study was conducted using data from the National Cancer Institute (NCI)-AIIMS, comprising 590 breast cancer cases and 1,366 controls. Data preprocessing included cleaning, missing value imputation, and feature engineering of 66 clinical, genetic, and lifestyle factors. To address class imbalance and multivariate complexities, the XGBoost ensemble model was employed. Model performance was evaluated using accuracy, recall, precision, F1-score, and AUC-ROC metrics. Gini index values were used to interpret model predictions and identify key features for risk stratification.

**Results:** The model demonstrated robust predictive performance with an accuracy of 0.89 and AUC-ROC > 0.9, sensitivity of 73.95%, and specificity of 94.90% on the test dataset. Feature importance analysis enabled the development of a reduced model using the top 20 features, maintaining high accuracy and clinical relevance. The reduced model simplifies risk assessment in resource-limited settings.

**Conclusions:** This pilot study introduces a population-specific ML-based breast cancer risk prediction tool tailored to the Indian demographic. By incorporating culturally relevant variables and leveraging advanced machine learning techniques, the model addresses key limitations of Western-centric risk prediction tools. While the current methodology serves as an initial framework, it can be further expanded and refined to develop more robust and generalizable models for broader population coverage. Integration into clinical workflows and further validation across diverse Indian populations could transform early detection and personalized intervention strategies, significantly reducing the burden of breast cancer in India.

## 1. Introduction

Breast cancer, the second most common cancer globally and the most frequent among women, accounted for over 2.3 million new cases and approximately 666,000 deaths worldwide in 2022.[1] The disease imposes a significant public health burden, particularly in low- and middle-income countries (LMICs), where limited healthcare infrastructure, low awareness, and socio-economic barriers contribute to delayed diagnosis and advanced-stage presentations. [2] Early detection and effective risk stratification are critical to improving survival rates, as early-stage breast cancer (e.g., Stage I) is associated with a significantly better prognosis compared to advanced stages (e.g., Stage III or IV). [3] Risk prediction models, such as the Gail Model, Tyrer-Cuzick Model (IBIS), BOADICEA, and BRCAPRO have been extensively used to identify individuals at high risk of developing breast cancer. These tools integrate multiple risk factors, including age, genetic predispositions, family history, hormonal exposures, and reproductive factors, to estimate breast cancer risk. [4,5] **Gail Model**, developed by Dr. Mitchell Gail and colleagues, evaluates factors such as age, reproductive history, prior breast biopsies, and ethnicity to estimate the risk of invasive breast cancer in women aged 35 years and older. [6,7] **The Tyrer-Cuzick Model** (IBIS Breast Cancer Risk Evaluation Tool) incorporates family history, genetic predispositions, hormonal influences, and lifestyle factors to provide a comprehensive breast cancer risk estimate, including probabilities for BRCA1 and BRCA2 mutations. [8,9] **The Claus Model** focuses on familial risk and genetic predispositions by analyzing detailed family history. **BRCAPRO**, developed at Duke University, considers family history, age of onset, and BRCA mutation testing results, making it particularly useful in genetic counseling. Similarly, the **CanRisk Tool**, formerly BOADICEA, integrates genetic, familial, and lifestyle factors to offer personalized risk estimates and is designed to support clinical genetics services. [10,11] Despite their widespread use, these models were predominantly developed and validated using data from Western populations, which limits their generalizability to non-Western settings. The Indian population presents unique challenges for breast cancer risk prediction. Genetic predispositions, such as the prevalence of BRCA1/BRCA2 mutations, vary significantly from those observed in Western populations. [12,13,14] Additionally, lifestyle and environmental factors such as dietary habits, reproductive behaviors, and healthcare access differ substantially. Cultural attitudes toward health and cancer screening, along with limited availability of genetic testing and detailed family history data, further complicate the application of Western risk models in the Indian context. [15] Consequently, these models often fail to provide accurate risk predictions, highlighting the need for tools tailored specifically to the Indian demographic. Machine learning (ML) offers a transformative approach to breast cancer risk prediction by leveraging its ability to analyze large, complex datasets and identify patterns that traditional statistical models might overlook.[16,17] ML-based models have demonstrated superior predictive accuracy compared to traditional models, with AU-ROC values exceeding 0.85 in certain studies.[18] By integrating diverse data types, including genetic, clinical, and lifestyle factors, ML models can adapt to population-specific characteristics and provide more personalized risk assessments. Furthermore, ML’s flexibility allows for the inclusion of emerging risk factors and the refinement of predictions over time.

Recognizing these challenges and opportunities, this pilot study aims to develop a machine learning–based tool for breast cancer risk assessment tailored to the Indian population. Utilizing a robust dataset comprising clinical, genetic, and lifestyle factors, the study addresses the limitations of existing models and aims to improve the accuracy of risk predictions. While this initial framework lays the groundwork, the methodology can be further expanded to develop more comprehensive and generalizable models for broader application. The proposed tool has the potential to facilitate early detection, enable targeted interventions, and enhance equity in breast cancer care across diverse Indian populations.

## 2. Materials and Methods

### 2.1 Study Design

This study was designed as a retrospective case-control pilot study to develop and validate a machine learning-based risk prediction model tailored to the Indian population. The study aimed to address the limitations of existing breast cancer risk prediction models by leveraging a robust dataset inclusive of genetic, clinical, and lifestyle factors.

### 2.2 Data collection Study Population

The data were obtained from the **National Cancer Institute (NCI)-AIIMS** and comprised:

- **Cases:** Women diagnosed with breast cancer (**n = 590**).
- **Controls:** Women without a breast cancer diagnosis (**n = 1,366**).

Upon receiving the dataset, a comprehensive data checks were conducted to identify inconsistencies, errors, and anomalies. This process involved checking for duplicate entries, outliers, missing values, and logical inconsistencies within the variables. Additionally, biological plausibility was assessed to ensure that all data points aligned with expected physiological and clinical patterns. Any discrepancies identified were either corrected through domain-informed imputations or removed to maintain the overall quality and reliability of the dataset for subsequent analysis.

Following a comprehensive data integrity assessment and the resolution of identified anomalies, the finalized dataset was prepared for modeling. This refined dataset ensured reliability and accuracy for subsequent predictive modeling.

#### Description of the Dataset

The dataset comprised 1,956 subjects with 66 features, covering a comprehensive range of demographic, medical, lifestyle, reproductive, and environmental factors. It was structured to facilitate multidimensional analysis and encompassed socioeconomic status, medical history, lifestyle choices, and physical attributes.

The dataset consisted of both numerical and categorical variables, each contributing to different aspects of analysis:

##### 1. Numerical Variables

These had included key demographic, health, reproductive, and physical measurement parameters such as Age, Age of Marriage, Duration of Marriage, Monthly Income, Age of Menarche, Age of Menopause (if applicable), Age if First Childbirth, Breastfeeding duration, Total number of Children, Total contraceptive use duration, Abortions (If applicable), Physical activity duration, Fruit vegetable intake, Weight, Height, Body mass index (BMI), and Waist -Hip ratio (WHR). These variables provided quantitative insights for statistical modelling, trend identification, and predictive analytics.

##### 2. Categorical Variables

Qualitative features included: Marital status, Education, Occupation, Residence, Socioeconomic Status (SES), Family type, Religion, Menstrual status, Regular Menstruation, Perimenopausal syndrome (Hot flushes), Breastfeeding, Breastfeeding category (complete, partial), Contraceptive, Contraceptive type, Abortion, Abortion type, Hormone replacement therapy, Family history of malignancy, Family history affected member type, Tobacco use, Tobacco type, Alcohol use, Dietary preferences (vegetarian/non vegetarian), Fasting, Fasting frequency, Fruit vegetable intake, Fat/Oil use, Oil type (mustard), History of Breast anomaly, Breast trauma, History of Radiation exposure, Radiation site (chest), Perspirant use (Perfume/Deodorant), and Talcum powder use etc. These had been essential for classification tasks, subgroup comparisons, and categorical trend analysis.

The dataset broadly consisted of the following domains:

- **Demographics:** The mean age was 47.87 (SD = 12.8) years (range: 18–87).
- **Medical History:** Prevalence of hypertension, diabetes, and thyroid disorders recorded.
- **Reproductive Health:** Mean menarche age was 14.45 (SD = 1.56) years, menopause age 46.15 (SD = 5.35) years, and breastfeeding duration 74.95 (SD = 46.68) months.
- **Genetic & Family History:** Cancer and hereditary conditions were documented.
- **Lifestyle Factors:** Data included tobacco/alcohol use, dietary habits, fasting, and physical activity (0–180 min).
- **Environmental Exposures:** Use of perfumes, deodorants, talcum powder, and radiation history recorded.
- **Anthropometrics:** Mean waist circumference was 87.59 (SD = 14.24) cm, hip circumference 97.35 (SD = 12.57) cm, with BMI variations across weight categories.

By distinguishing between numerical and categorical variables, the dataset supported a wide range of analytical approaches, from statistical modelling and correlation analysis to feature selection and predictive modelling applications. A few variables representing the demographic, reproductive, lifestyle, environment and health-related variables were visualized such as menstrual status, distribution of age, family history of cancer, distribution of menarche age, abortion frequency and distribution of residence, distribution of tobacco use, physical activity grade etc. as shown in the figure 1.

**Fig 1.**
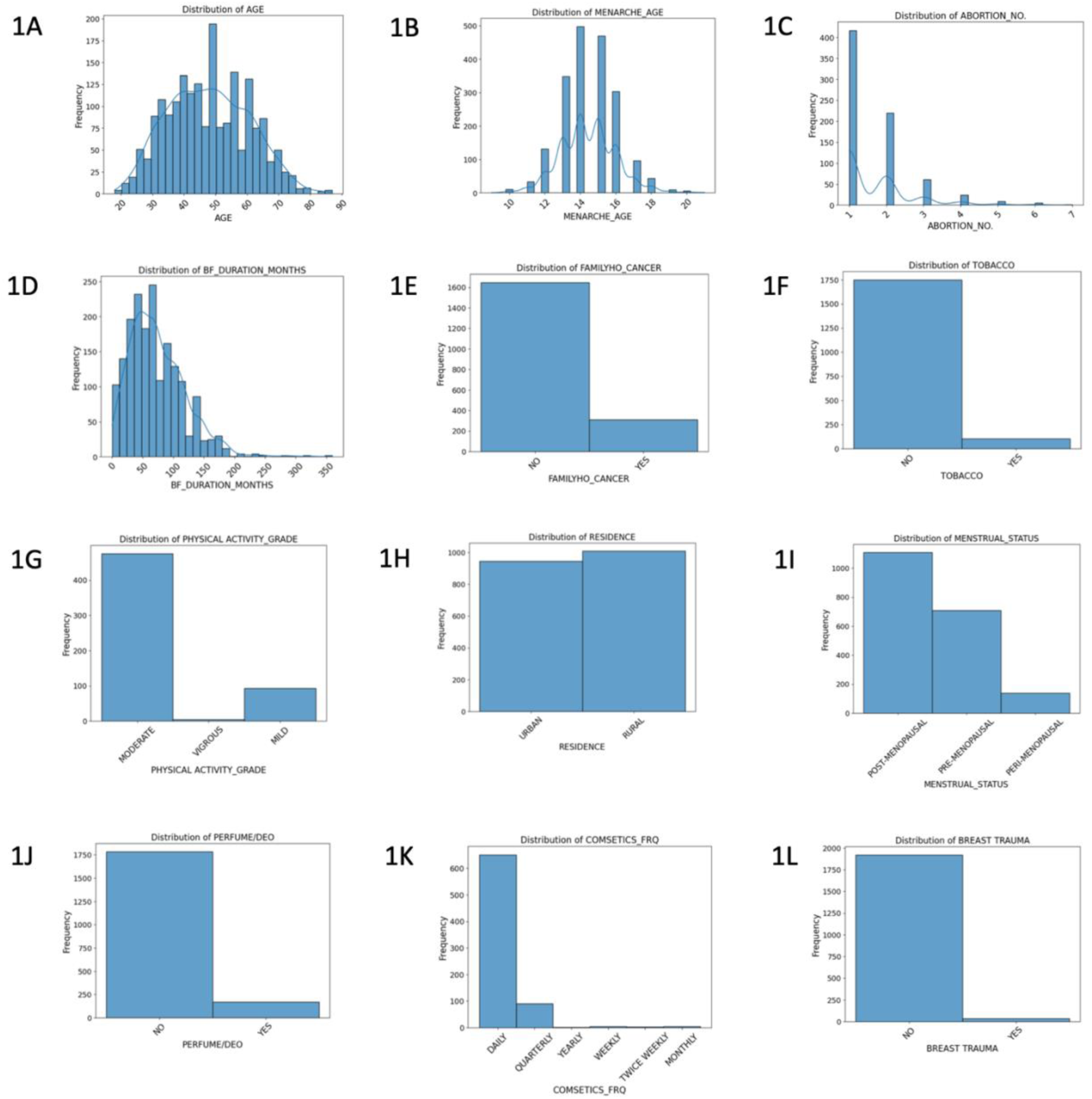
(A-L): This figure presents the distribution of some of the demographic, reproductive, lifestyle, environment and health-related variables in the dataset. Each histogram depicts the frequency of subjects across different categories or numerical values

### 2.3 Data Splitting and Data Preprocessing

The table 1 presents the distribution of cases and controls used in the modelling strategy. The dataset included a total of 590 cases and 1,366 controls.

**Table 1.**
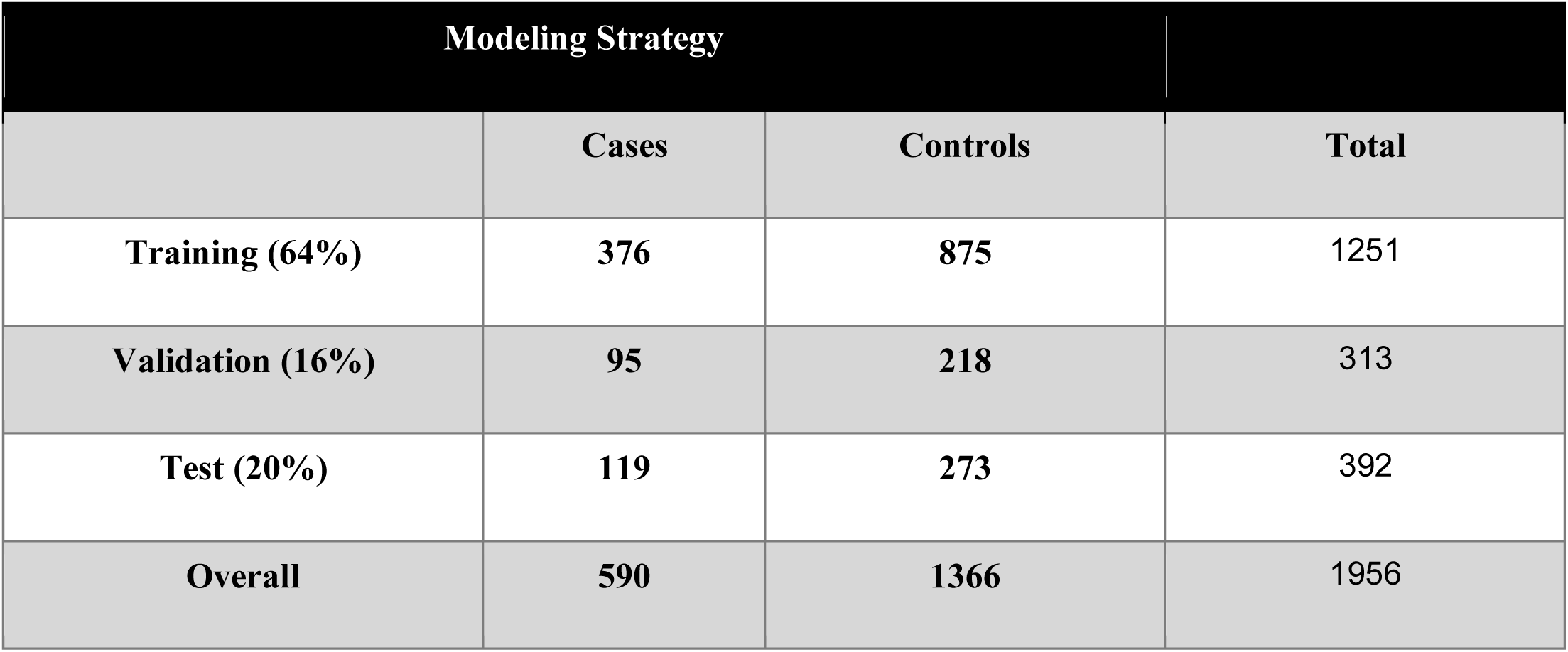
Distribution of Cases and Controls for Modeling Strategy.

Prior to data pre-processing, 20% of the raw dataset was reserved as an unseen test set to mitigate the risk of data leakage during model evaluation. The remaining 80% was subsequently partitioned into a training set and a validation set, with the latter, utilized for hyperparameter tuning.

Rigorous data pre-processing is essential for improving model performance, ensuring accuracy, and enhancing reliability. The pre-processing workflow encompassed several critical steps, including data wrangling, data cleaning, missing value imputation, and feature engineering, to optimize the dataset for machine learning applications. Data wrangling involved the integration of multiple datasets into a unified structure while maintaining consistency in format and structure. This was followed by data cleaning to address inconsistencies and handle missing values, thereby ensuring the dataset was well-prepared for modeling.

The prepared dataset contained a substantial number of missing values. Given that certain missing values may hold biological significance, a threshold of 65% was established based on the proportion of missing values across categorical and numerical variables. Data points exceeding this threshold were excluded from the analysis to maintain data integrity and ensure meaningful imputation.

To address the remaining missing values while preserving biological relevance, **three distinct imputation strategies** were employed:

1. Not Applicable (NA) Imputation:

○ Features such as **Age of Marriage, Duration of Marriage, Age if First Childbirth, Breastfeeding duration, Total number of Children, Total Contraceptive use duration, Total number of Abortions** were imputed with zero (**0)** to represent “Not Applicable” scenarios.
○ For instance, women who were never married would have missing values for **Age of Marriage** and **Duration of Marriage**, as these variables were irrelevant to their profiles.
2. Most Frequent Value Imputation:

○ Variables such as **Age, Age of Menarche, Monthly Income, Physical activity duration, Weight, Height, Hip circumference, Waist circumference and Marital status** were imputed using their **mode (most frequent value)** to retain population-level trends.
3. Menopause-Related Imputation:

○ Missing values in **Age of Menopause** were imputed based on menopausal status:

▪ **Pre-menopausal females** → Imputed with **-1** (Not Applicable).
▪ **Peri-menopausal females** → Imputed with **0**.
▪ **Post-menopausal females** → Imputed with the **most frequent value** (46).

These imputation strategies were carefully designed to minimize data loss while preserving the biological significance of missing values, ensuring that imputations were biologically meaningful and aligned with the overall data structure.

Subsequently, **Feature Engineering** was performed. Numerical variables were scaled using the **Min-Max Scaler**, as the dataset had no significant outliers that could influence the scaling process. Since biological data integrity was ensured during preprocessing, potential outliers had already been addressed, making **Min-Max Scaling** a suitable choice for normalization. This method preserved the original distribution of the data while rescaling features to a standardized range, ensuring consistency across numerical variables.

**Min-Max Scaling** was preferred over **Standard Scaling** due to the presence of both **categorical and numerical features** in the dataset. **Standard Scaler (Z-score normalization)** transforms data to have a mean of zero and a standard deviation of one, which assumes a normal distribution. However, this assumption does not always hold for biomedical data, where feature distributions can vary significantly. In contrast, **Min-Max Scaling** scales all numerical features to a fixed range (typically [0,1]), making it more interpretable and compatible with models that are sensitive to varying feature magnitudes. Additionally, since frequency encoding was applied to categorical variables, using **Min-Max Scaling** ensured that numerical features remained on a similar scale, preventing any imbalance that could arise from mixed feature types.

Categorical variables were encoded using **frequency encoding** to efficiently handle high-cardinality features. In this approach, each category was replaced by its occurrence frequency in the respective column. **Frequency encoding** was chosen over **one-hot encoding** to avoid the curse of dimensionality, reduce memory usage, and maintain computational efficiency, particularly given the presence of categorical variables with numerous unique values.

Upon completion of the data preprocessing pipeline, the final dataset comprised **471 cases (patients with breast cancer) and 1,093 controls (individuals without breast cancer), with no missing values**. The dataset was **multivariate, consisting of 66 features per data point**, ensuring a comprehensive and structured input for predictive modeling. The modelling strategy can be summarized as follows:

### 2.4 Modelling strategy

#### 2.4.1 Risk Prediction Model Training

The dataset exhibited a significant class imbalance, with the number of patients being substantially lower than the number of controls, leading to a skewed distribution. Standard machine learning algorithms often struggle with such imbalanced datasets, as they tend to be biased toward the majority class, reducing prediction accuracy for the minority class. To address this challenge, an ensemble learning approach was employed, leveraging multiple base classifiers to enhance predictive performance. Given its efficiency in handling high-dimensional, multivariate data, XGBoost (Extreme Gradient Boosting) was chosen as the primary model for risk prediction. Unlike traditional decision trees that are prone to overfitting, XGBoost sequentially builds trees, with each tree correcting the errors of its predecessors, thereby improving predictive accuracy while maintaining model generalizability. Additionally, XGBoost incorporates several key mechanisms to enhance performance. Instead of randomly selecting features at each split, it chooses the most informative feature based on splitting criteria such as information gain. The colsample_bytree parameter, set to 0.8, introduced randomness by selecting a subset of features per tree, thereby reducing the risk of overfitting. To further address class imbalance, the Adaptive Synthetic (ADASYN) sampling technique was applied to generate synthetic samples for the minority class, ensuring a balanced representation within the training set. Although XGBoost does not explicitly decorrelate features, its gradient boosting framework iteratively reduces errors, allowing the model to effectively capture complex feature interactions. Furthermore, XGBoost employs various techniques to prevent overfitting, including shrinkage (learning rate control), L1/L2 regularization, and subsampling. These mechanisms make the model resilient to outliers and noise, which are common challenges in high-dimensional biomedical datasets. Given the complexity of biomedical data, where multiple features interact, XGBoost efficiently captures these relationships through optimized decision splits, making it a robust tool for breast cancer risk prediction.

Prior to training, the dataset was split into training and validation sets, with the validation set used for hyperparameter tuning, while a separate test set was reserved as an unseen dataset for final evaluation. The training dataset contained 1,251 samples, while the validation dataset comprised 313 samples. ADASYN was applied to generate synthetic samples for the minority class, ensuring a balanced training dataset. Following this, the XGBoost model was trained using the scikit-learn Python package. The model was optimized using hyperparameter tuning, with max_depth set to 6 to prevent excessively complex trees and eta (learning rate) set to 0.2 to control the step size during training. The subsample parameter was set to 1 to ensure all data points were utilized during training, while colsample_bytree was set to 0.8 to introduce feature selection randomness. The objective function binary:logistic was selected to suit the classification task, with logloss used as the evaluation metric. Early stopping with 10 rounds was implemented to halt training if no improvement was observed, preventing unnecessary computations. Additionally, num_boost_round was set to 100 to control the number of boosting iterations, ensuring sufficient model training, while nfold was set to 3 to facilitate cross-validation and assess model performance. A seed value of 42 was used to ensure reproducibility.

This carefully optimized XGBoost model provided a robust, well-generalized framework for predicting breast cancer risk while effectively handling class imbalance as depicted in Table 2. The combination of ensemble learning, gradient boosting, and hyperparameter tuning allowed the model to capture complex feature interactions, improving both predictive accuracy and reliability in a high-dimensional biomedical dataset.

**Table 2.** Overview of the XGBoost Modeling Strategy and Hyperparameters.

| Modeling Strategy (XGBoost) |  |
| --- | --- |
| Overall Dataset | 1956 subjects (590 Breast Cancer patients + 1366 controls) |
| Training Dataset | 1251 subjects ( 376 Breast Cancer patients + 875 controls) |
| Validation Dataset | 313 subjects ( 95 Breast Cancer patients + 218 controls) |
| Hyperparameters | Values |
| max_depth | 6 |
| eta | 0.2 |
| subsample | 1 |
| colsample_bytree | 0.8 |
| objective | binary:logistic |
| num_boost_round | 100 |
| nfold | 3 |
| eval_metric | logloss |
| early_stopping_rounds | 10 |
| seed | 42 |
| class_weight | ADASYN for class balancing |

#### 2.4.2. Evaluation methodology

The XGBoost model was trained after applying class balancing techniques and subsequently evaluated on an unseen test dataset using various performance metrics, including Accuracy, Recall, Precision, and F1-Score, along with the Receiver Operating Characteristic (ROC) and Precision-Recall (P-R) curves. These metrics are defined in terms of True Positives (TP), False Positives (FP), True Negatives (TN), and False Negatives (FN) for a given class as:

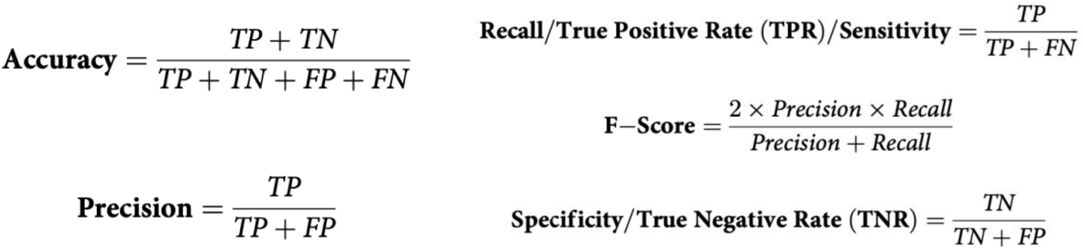

Accuracy represents the proportion of correctly classified instances, Recall (Sensitivity) measures the ability to identify positive cases, Precision quantifies the proportion of predicted positive instances that are actually positive, and the F1-Score is the harmonic mean of Precision and Recall, providing a balanced evaluation of model performance.

The ROC curve plots True Positive Rate (TPR) against False Positive Rate (FPR = 1 - Specificity or True Negative Rate), illustrating the model’s ability to differentiate between classes. The Area Under the Curve (AUC-ROC) quantifies overall classification performance, with higher values indicating better discrimination between positive and negative cases. The sklearn.metrics module was employed to compute accuracy_score(), recall_score(), precision_score(), and f1_score(), while plot_roc_curve() and precision_recall_curve() were used to visualize the ROC and P-R curves and derive their respective AUC values.

To identify the most important features contributing to breast cancer risk prediction, the Gini index was used. The Gini index measures the impurity reduction achieved by each feature at decision tree splits within the XGBoost model. Features with higher Gini scores had a greater impact on model decisions, allowing for the extraction of the top predictive factors in the dataset.

## 3. Ethical Considerations

Approval for this study was obtained from the Institutional Ethics Committee at AIIMS, New Delhi (approval number: [IEC: 499/17.06.2022, RP-01/2022]). Written informed consent was obtained from all participants, ensuring confidentiality and voluntary participation. No patient could be re-identified from the data used in this study. The study deidentified patient records and did not involve the use, or transmittal of individually identifiable data

## 4. Results

The XGBoost model was trained on a dataset comprising 1,251 subjects, including 376 breast cancer patients and 875 controls. The model’s performance was first evaluated on a validation dataset consisting of 313 subjects (95 breast cancer patients and 218 controls) before being tested on an unseen test dataset of 392 subjects (119 breast cancer patients and 273 controls).

### 4.1 Evaluation of the Breast Cancer Risk Prediction Model

Performance metrics for the breast cancer risk prediction model are given in the table 3.

**Table 3:** Performance of Breast Cancer Prediction Model.

|  |  |
| --- | --- |
| **Training Dataset** | **1251 subjects (376 Breast Cancer patients + 875 controls)** |
| **Validation Dataset** | **313 subjects (95 Breast Cancer patients + 218 controls)** |
| **Training Accuracy on Validation Dataset** | **0.89** |
| **Validation Dataset F1-Score** | **0.92(for class control (0))**<br>**0.80(for class cancer (1))** |
|  | **Macro F score:0.86** |
| **Test Dataset** | **392 subjects (119 Breast Cancer patients + 273 controls)** |
| **Test Dataset Accuracy** | **0.89** |
| **Mean F1-Score on Test Dataset** | **0.92(for class control (0))**<br>**0.80(for class cancer (1))**<br>**Macro F score:0.86** |
| **Sensitivity (Recall) on Test Dataset** | **0.7395** |
| **Specificity on Test Dataset** | **0.949** |
| **PPV (Precision) on Test Dataset** | **0.8627** |
| **NPV on Test Dataset** | **0.8931** |
| **AUC-ROC (on Test Dataset)** | **0.92** |
| **AUC-PRC (on Test Dataset)** | **0.89** |

On the validation dataset, the model achieved an overall accuracy of 0.89. The F1-score for the control class (0) was 0.92, while the F1-score for the cancer class (1) was 0.80, resulting in a macro F-score of 0.86. These results demonstrated the model’s ability to distinguish between cancer and control cases with a strong balance between precision and recall.

On the final test dataset, the model maintained an accuracy of 0.89, confirming its generalizability. The mean F1-score remained consistent with the validation results, with a value of 0.92 for the control class and 0.80 for the cancer class, yielding a macro-F-score of 0.86. The model’s sensitivity (recall) on the test dataset was 0.7395, indicating that approximately 73.95% of actual cancer cases were correctly identified. The specificity was 0.949, reflecting the model’s ability to correctly classify non-cancer cases. Additionally, the Positive Predictive Value (PPV or precision) was 0.8627, signifying that 86.27% of the predicted cancer cases were truly cancerous, while the Negative Predictive Value (NPV) was 0.8931, indicating that 89.31% of predicted controls were actual controls.

The model’s overall discriminative ability was further validated through the AUC-ROC (Area Under the Receiver Operating Characteristic Curve), which was 0.92, demonstrating strong separation between the two classes. The AUC-PRC (Area Under the Precision-Recall Curve) was 0.89, confirming the model’s robustness in handling class imbalances while maintaining high predictive accuracy. The above results, as summarised and depicted in Figure 2 and Table 3, indicate that the XGBoost model provides a reliable and well-balanced approach to breast cancer risk prediction. Figure 3 gives a graphical representation of the decision tree used in the XGBoost model, illustrating the hierarchical structure of feature-based decision splits. Each node represents a decision based on a specific feature, with branches indicating the corresponding conditions. Blue and red arrows denote different decision paths, leading to classification outcomes. The depth and complexity of the tree highlight key predictive variables influencing breast cancer classification. The tree starts with the most important feature that best splits the dataset into different groups. For example, this could be **age at first childbirth**, **family history of breast cancer**, or **menstrual history**.

**Figure 2.**
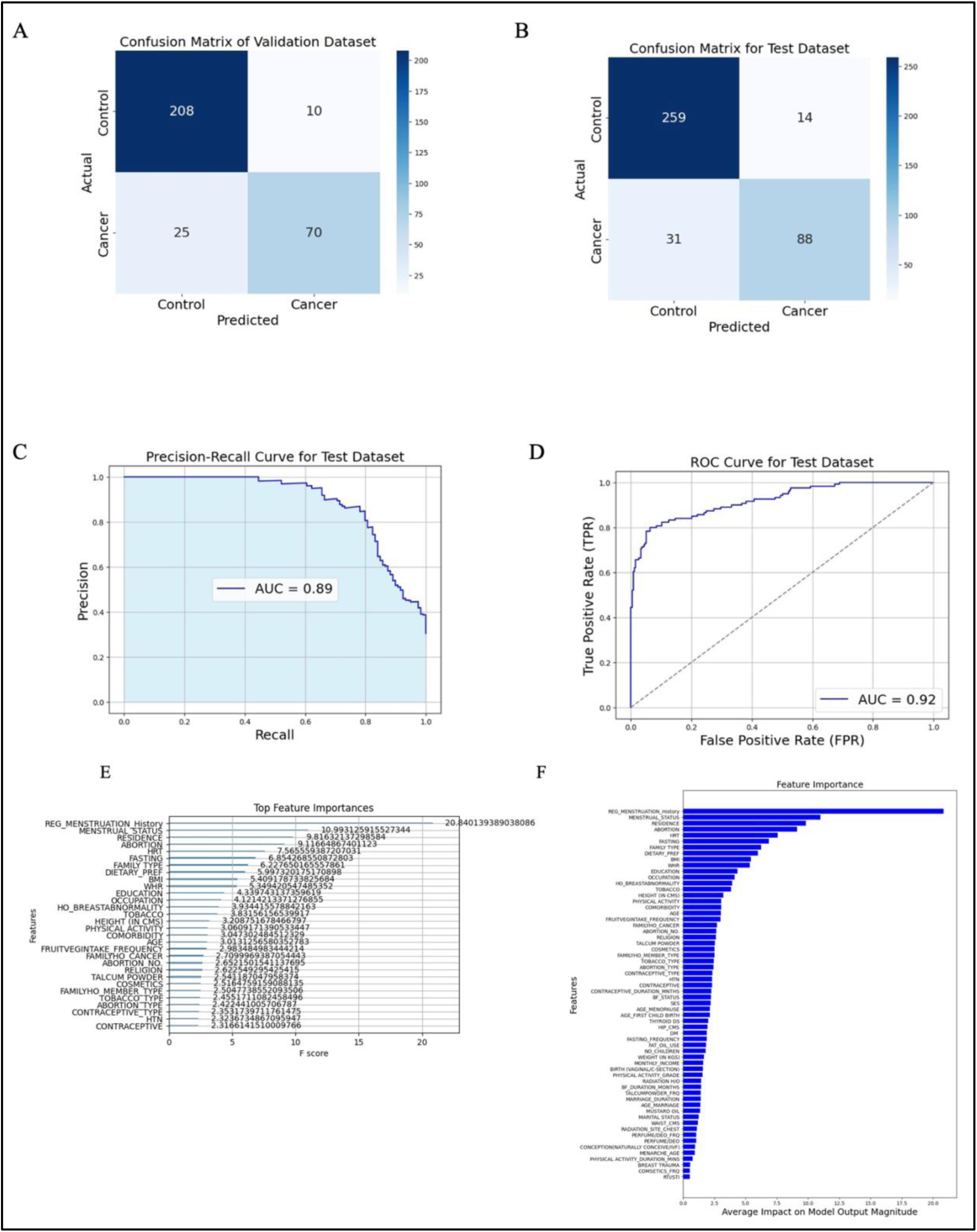
Performance Evaluation and Feature Importance of the XGBoost Model: (A, B) Confusion matrices for the validation and test datasets, respectively, showing true positives, true negatives, false positives, and false negatives. The model demonstrates high classification performance, with minimal misclassifications in both datasets. (C) Precision-recall (PR) curve for the test dataset, with an area under the curve (AUC) of 0.89, indicating strong predictive capability, particularly in handling class imbalance. (D) Receiver operating characteristic (ROC) curve for the test dataset, with an AUC of 0.92, highlighting the model’s robust discrimination between breast cancer cases and controls. (E, F) Feature importance analysis using the Gini index, displaying the most influential predictors in the model. The highest-ranked features include menstrual history, menstrual status, abortion frequency, and family history of cancer, among others.

**Figure 3.**
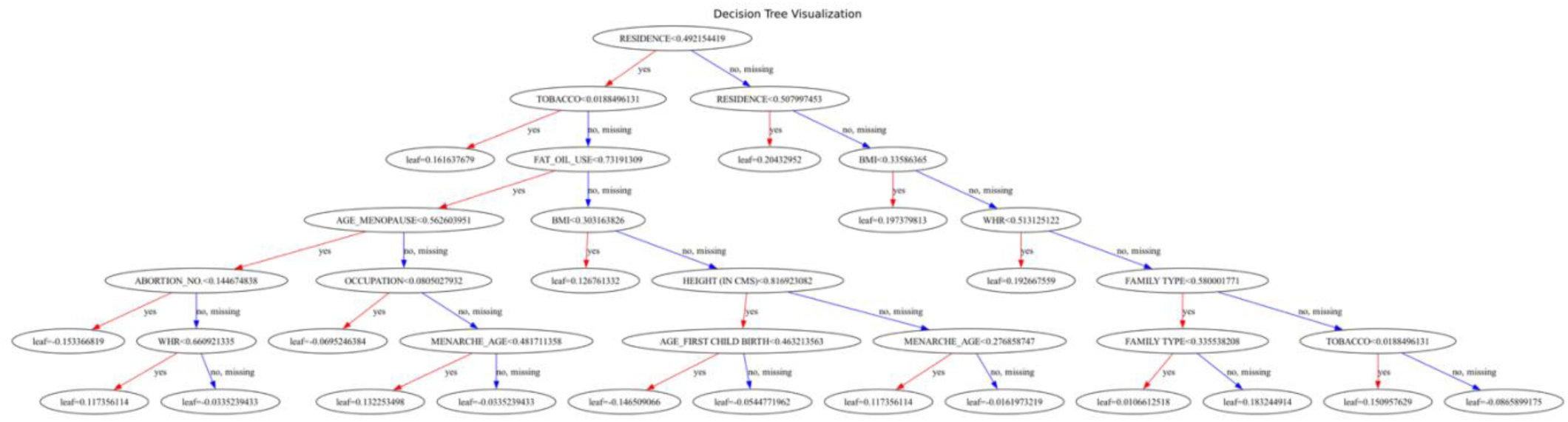
Decision Tree Visualization

**Figure 4.**
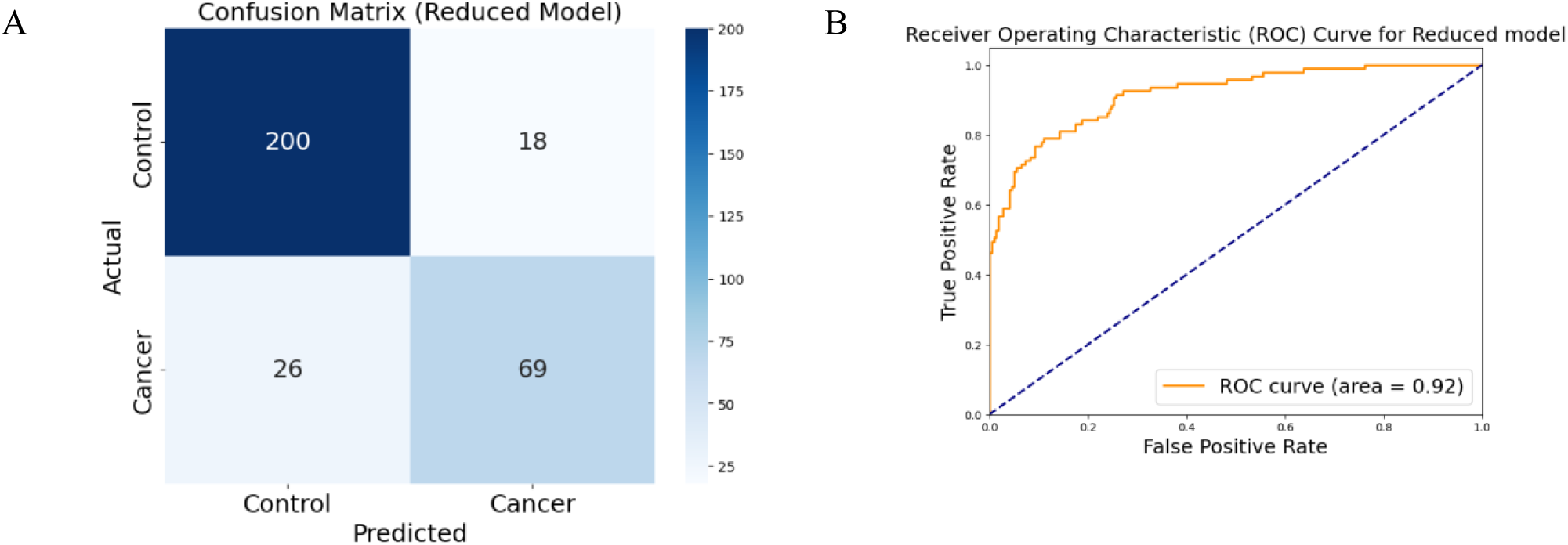
Performance Evaluation of the Reduced XGBoost Model: A) **Confusion Matrix**: The confusion matrix for the reduced model shows the classification performance on the test dataset. (B) **Receiver Operating Characteristic (ROC) Curve**: The ROC curve evaluates the model’s ability to distinguish between cancer and control cases. The area under the curve (AUC) is 0.92, indicating strong predictive performance. The solid orange line represents the model’s performance, while the dashed blue line represents random chance (AUC = 0.5).

Each node represents a feature-based condition. If the condition is met (e.g., “Age at first childbirth < 30”), the decision moves to the left; otherwise, it moves to the right. The tree continues splitting based on different features that help refine predictions based on known risk factors. The endpoints (leaf nodes) indicate the final prediction, which could be **high risk (cancer) or low risk (control)**. The probability of developing cancer can also be represented at each leaf node.

### 4.2 Reduced models

Our machine learning models utilize 66 clinical parameters for breast cancer risk prediction. However, in many real-world clinical settings, acquiring all 66 features may not be feasible due to time constraints or limited availability of certain patient data. To address this challenge, we analyzed the feature importance rankings derived from an XGBoost-trained model and identified the top 20 most influential features. These features were selected based on their contributions to model performance.

To develop a more practical and efficient predictive model, we constructed a “reduced” version that utilizes only these top 20 features. This reduction in input variables allows for faster and more accessible data collection while maintaining predictive performance. Following feature selection, we re-optimized the model by conducting hyperparameter tuning specifically for the reduced feature set.

Despite the significant reduction in the number of input features, the optimized reduced model demonstrated strong predictive performance. On the validation set, it achieved an accuracy of 0.86 and an AUC-ROC score of 0.92, which are comparable to the original full-feature model. These results indicate that the reduced model effectively balances efficiency and predictive power, making it a viable option for clinical implementation where access to all 66 parameters may be impractical.

## 5. Discussion

While the existing breast cancer risk prediction calculators have been instrumental in early detection and prevention efforts in Western countries, their applicability in the Indian context is limited due to demographic, genetic, lifestyle, and cultural differences. There was a need for developing and validating breast cancer risk prediction models tailored specifically to the Indian population to improve their accuracy and utility in this context.

Following this we endeavored to build a supervised machine learning model pertaining to the needs of the Indian population. This study introduces a machine learning (ML)--based breast cancer risk prediction model uniquely tailored to the Indian population, addressing critical gaps in traditional risk prediction tools. Models such as Gail, Tyrer-Cuzick, and BOADICEA, though instrumental in early detection efforts globally, were developed using predominantly Western datasets and fail to account for the unique socio-demographic, genetic, and lifestyle factors prevalent in India. For instance, the prevalence of BRCA mutations, cultural barriers to screening, and dietary habits differ significantly between Indian and Western populations, impacting the accuracy of these models in non-Western contexts. [19,20,21] The Gail model, in particular, demonstrates low sensitivity in Indian settings due to its reliance on risk factors such as nulliparity, late age at first childbirth, and hormone replacement therapy, which are less relevant to Indian women. [22]. A hospital-based study evaluating the Gail model in Western India revealed a sensitivity of only 10.3% and highlighted its underperformance in accurately stratifying Indian women by risk, underscoring the need for population-specific risk assessment tools. [23]

Machine learning-based models, such as the XGBoost model used in this study have demonstrated the superior performance of an ensemble-based model, achieving a high predictive accuracy (AUC-ROC > 0.9). Unlike the static approach of traditional models, ensemble learning techniques are adept at handling complex, multivariate datasets, as demonstrated in previous studies.[24] This adaptability allows for the inclusion of culturally relevant variables and improves the model’s relevance and predictive accuracy for the Indian population.

One of the strengths of this study lies in addressing the inherent class imbalance in the dataset, which included 590 cases and 1,366 controls, a common challenge in breast cancer datasets. The application of ADASYN (Adaptive Synthetic Sampling) improved class balance, leading to a recall of 74% and a specificity of 95% on the test dataset. Feature importance was determined using the Gini index, enabling the selection of key predictors that contributed most to classification performance. This facilitated the development of a reduced model using the top-ranked features, maintaining high accuracy while improving clinical applicability. Such reduced models are particularly valuable in resource-limited settings, where comprehensive data collection may not always be feasible.

Unlike static, regression-based methods, ML models can dynamically incorporate emerging risk factors, making them more adaptable and robust for use in diverse populations. For instance, the inclusion of Indian-specific factors in our model addresses the global discourse on the limitations of Western-developed models in capturing gene-environment interactions unique to Asian populations.[23] However, despite these advancements, significant challenges remain in implementing ML models effectively in low- and middle-income countries (LMICs), including limited access to data, infrastructural constraints, and disparities in healthcare delivery.

This pilot study provides a significant step toward developing accessible and effective breast cancer risk prediction tools for the Indian population, with implications for early detection and targeted interventions. By integrating such models into clinical workflows, healthcare providers could enhance screening efficiency and personalized risk management strategies, ultimately reducing the burden of advanced-stage breast cancer in India.

## 6. Recommendations

The high predictive accuracy of our model highlights its potential utility as a clinical screening tool.

However, for successful implementation, several considerations must be addressed:

1. **External Validation**: external validation across diverse Indian populations is essential to confirm generalizability and robustness. Multi-center studies involving varied demographic and socio-economic groups would ensure the model’s applicability across India.
2. **Integration with Healthcare Systems:** To facilitate clinical adoption, the development of user-friendly interfaces and seamless integration with existing healthcare workflows are crucial.
3. **Continuous Learning:** Mechanisms for incorporating new data into the model are vital for sustained relevance and accuracy. A continuous learning framework, where the model is periodically updated with data from different populations and evolving risk factors, would enhance its clinical utility.
4. **Training for Healthcare Providers**: Effective use of the model requires educating healthcare providers on its applications and limitations. Training programs can help ensure accurate interpretation of risk scores and guide appropriate clinical decision-making.
5. **Policy-Level Support:** To maximize the model’s impact, collaboration with public health authorities to integrate it into national cancer control programs is essential. Risk prediction tools could complement existing screening programs by identifying high-risk individuals for targeted screening and risk reduction interventions.

## 7. Limitations of the Study

1. **Retrospective Design:** The study uses retrospective data, which may not fully capture the temporal and evolving nature of breast cancer risk factors, potentially limiting its ability to predict future trends.
2. **Selection Bias:** The dataset was derived from a single institution, which may introduce selection bias and limit the generalizability of the findings to broader, diverse populations across India.
3. **Limited External Validation:** While the model demonstrated strong internal validation, its performance has not yet been validated on external datasets or populations. This is necessary to confirm its robustness and applicability.
4. **Cost and Accessibility:** The application of machine learning models in routine clinical settings may face challenges related to cost, infrastructure, and digital literacy, particularly in rural and underserved areas.

## 8. Conclusion

This study represents a significant advancement in developing a population-specific breast cancer risk prediction model for Indian women. By addressing the limitations of traditional tools and leveraging machine learning, the model provides a foundation for personalized risk assessment and targeted early interventions. Successful clinical implementation, combined with further validation and refinements, has the potential to transform breast cancer prevention and care in India, reducing the burden of advanced-stage diagnoses and contributing to more equitable healthcare outcomes.

## Data availability statement

The data supporting the conclusions of this study can be obtained upon request from Dr Jitendra Kumar Meena, the corresponding author. However, the data cannot be made publicly accessible due to its containing information that might jeopardize the privacy of the research participants.

## Acknowledgment

The authors would like to thank all the participants who agreed to take part in the study. Acknowledgement. We acknowledge and thank the funding support from Ministry of Education, Government of India, Central Project Management Unit, IIT Jammu with sanction number IITJMU/CPMU-AI/2024/0002.

## References

1. Bray F, Laversanne M, Sung H, Ferlay J, Siegel RL, Soerjomataram I, Jemal A. Global cancer statistics 2022: GLOBOCAN estimates of incidence and mortality worldwide for 36 cancers in 185 countries. CA: a cancer journal for clinicians. 2024 May;74(3):229–63.

2. Weerarathna IN, Luharia A, Uke A, Mishra G. Challenges and Innovations in Breast Cancer Screening in India: A Review of Epidemiological Trends and Diagnostic Strategies. International Journal of Breast Cancer. 2024;2024(1):6845966.

3. Ginsburg O, Yip CH, Brooks A, Cabanes A, Caleffi M, Dunstan Yataco JA, Gyawali B, McCormack V, McLaughlin de Anderson M, Mehrotra R, Mohar A. Breast cancer early detection: A phased approach to implementation. Cancer. 2020 May 15;126:2379–93.

4. Cintolo-Gonzalez JA, Braun D, Blackford AL, Mazzola E, Acar A, Plichta JK, Griffin M, Hughes KS. Breast cancer risk models: a comprehensive overview of existing models, validation, and clinical applications. Breast cancer research and treatment. 2017 Jul;164:263–84.

5. Antoniou AC, Hardy R, Walker L, Evans DG, Shenton A, Eeles R, Shanley S, Pichert G, Izatt L, Rose S, Douglas F. Predicting the likelihood of carrying a BRCA1 or BRCA2 mutation: validation of BOADICEA, BRCAPRO, IBIS, Myriad and the Manchester scoring system using data from UK genetics clinics. Journal of medical genetics. 2008 Jul 1;45(7):425–31.

6. Thomas S, Suhani, Desai G, Pathania OP, Jain M, Aggarwal L, Ali S, Sharma K, Tudu SK. Clinico-epidemiological profile of breast cancer patients and the retrospective application of Gail model 2: an Indian perspective. Breast Disease. 2016 Mar 28;36(1):15–22.

7. Stevanato KP, Pedroso RB, Agnolo CM, Dos Santos L, Pelloso FC, de Barros Carvalho MD, Pelloso SM. Use and applicability of the Gail model to calculate breast cancer risk: a scoping review. Asian Pacific Journal of Cancer Prevention: APJCP. 2022 Apr;23(4):1117.

8. Brentnall AR, Cuzick J. Risk models for breast cancer and their validation. Statistical science: a review journal of the Institute of Mathematical Statistics. 2020 Mar 3;35(1):14.

9. Kurian AW, Hughes E, Simmons T, Bernhisel R, Probst B, Meek S, Caswell-Jin JL, John EM, Lanchbury JS, Slavin TP, Wagner S. Performance of the IBIS/Tyrer-Cuzick model of breast cancer risk by race and ethnicity in the Women’s Health Initiative. Cancer. 2021 Oct 15;127(20):3742–50.

10. Berry DA, Iversen Jr ES, Gudbjartsson DF, Hiller EH, Garber JE, Peshkin BN, Lerman C, Watson P, Lynch HT, Hilsenbeck SG, Rubinstein WS. BRCAPRO validation, sensitivity of genetic testing of BRCA1/BRCA2, and prevalence of other breast cancer susceptibility genes. Journal of Clinical Oncology. 2002 Jun 1;20(11):2701–12.

11. Jacobi CE, de Bock GH, Siegerink B, van Asperen CJ. Differences and similarities in breast cancer risk assessment models in clinical practice: which model to choose?. Breast cancer research and treatment. 2009 May;115:381–90.

12. Singh J, Thota N, Singh S, Padhi S, Mohan P, Deshwal S, Sur S, Ghosh M, Agarwal A, Sarin R, Ahmed R. Screening of over 1000 Indian patients with breast and/or ovarian cancer with a multi-gene panel: prevalence of BRCA1/2 and non-BRCA mutations. Breast cancer research and treatment. 2018 Jul;170:189–96.

13. Kim H, Choi DH. Distribution of BRCA1 and BRCA2 mutations in Asian patients with breast cancer. Journal of breast cancer. 2013 Dec;16(4):357.

14. Patra A, Ali SS, Devi NM, Qadeer AS, Kamalakannan S, Nag S, Kulkarni SS, Rajappa S, Hariharan N, Pant HB, Agiwal V. Prevalence of BRCA mutation in breast and ovarian cancer among women in India: A systematic review and meta-analysis protocol. Plos one. 2024 Jul 16;19(7):e0306612.

15. Bhardwaj PV, Dulala R, Rajappa S, Loke C. Breast cancer in India: screening, detection, and management. Hematology/Oncology Clinics. 2024 Feb 1;38(1):123–35.

16. Ming C, Viassolo V, Probst-Hensch N, Chappuis PO, Dinov ID, Katapodi MC. Machine learning techniques for personalized breast cancer risk prediction: comparison with the BCRAT and BOADICEA models. Breast Cancer Research. 2019 Dec;21:1–1.

17. Stark GF, Hart GR, Nartowt BJ, Deng J. Predicting breast cancer risk using personal health data and machine learning models. Plos one. 2019 Dec 27;14(12):e0226765.

18. Hussain S, Ali M, Naseem U, Nezhadmoghadam F, Jatoi MA, Gulliver TA, Tamez-Peña JG. Breast cancer risk prediction using machine learning: a systematic review. Frontiers in Oncology. 2024 Mar 20;14:1343627.

19. Leong SP, Shen ZZ, Liu TJ, Agarwal G, Tajima T, Paik NS, Sandelin K, Derossis A, Cody H, Foulkes WD. Is breast cancer the same disease in Asian and Western countries?. World journal of surgery. 2010 Oct;34:2308–24.

20. Fackenthal JD, Olopade OI. Breast cancer risk associated with BRCA1 and BRCA2 in diverse populations. Nature Reviews Cancer. 2007 Dec;7(12):937–48.

21. Lila K, Bhanushali H, Chanekar M, Jatale R, Banerjee M, Dixit RB, Rajadhyaksha A, Chadha K. Mutation Spectrum Analysis of BRCA1/2 Genes for Hereditary Breast and Ovarian Cancer in the Indian Population. Asian Pacific Journal of Cancer Prevention. 2024 Dec 1;25(12):4145–51.

22. Thomas S, Suhani, Desai G, Pathania OP, Jain M, Aggarwal L, Ali S, Sharma K, Tudu SK. Clinico-epidemiological profile of breast cancer patients and the retrospective application of Gail model 2: an Indian perspective. Breast Disease. 2016 Mar 28;36(1):15–22.

23. Kumar N, Singh V, Mehta G. Assessment of common risk factors and validation of the Gail model for breast cancer: A hospital-based study from Western India. Tzu Chi Medical Journal. 2020 Oct 1;32(4):362–6.

24. Jose R, Augustine P, Paul L, Haran JC, Subramanian S. Development and validation of the Snehita BRISK model: A breast cancer risk assessment tool for risk stratification in women of the Indian subcontinent. Clinical Epidemiology and Global Health. 2025 Jan 1;31:101884.

